# Projecting the combined healthcare burden of seasonal influenza and COVID-19

**DOI:** 10.1101/2020.12.20.20248599

**Authors:** Zhanwei Du, Spencer J. Fox, Tanvi Ingle, Michael P. Pignone, Lauren Ancel Meyers

## Abstract

The overlapping 2020-2021 influenza season and COVID-19 pandemic may overwhelm hospitals throughout the Northern Hemisphere. Using a mathematical model, we project that COVID-19 burden will dwarf that of influenza. If non-pharmacological mitigation efforts fail, increasing influenza vaccination coverage by 30% points would avert 54 hospitalizations per 100,000 people.

---

As of November 2020, the COVID-19 pandemic has surpassed eleven million confirmed cases and 240,000 deaths in the United States (US). The imminent emergence of seasonal influenza in the Northern Hemisphere risks overwhelming already strained healthcare systems, particularly if influenza immunization rates are low. However, data from the Southern Hemisphere (1) and the 2019-2020 influenza season (2) suggest that COVID-19 mitigation efforts may suppress the spread of influenza. Here, we use a data-driven model of the co-circulation of influenza and SARS-CoV-2 to project the combined healthcare burden of the 2020-2021 influenza season and COVID-19 pandemic under different levels of non-pharmacological interventions and influenza immunization rates.

Our deterministic susceptible-exposed-infected-recovered (SEIR) compartmental model incorporates age-specific viral transmission rates and disease severity (Figure S5). Extending prior policy analyses (3) for Travis County, which contains most of Austin, Texas, we model three observed levels of SARS-CoV-2 transmission and assume that mitigation equally impacts both viruses: low (late May, after the relaxation of stay-home orders), medium (mid June, summer wave), and high (early March, prior to all measures) (Table S1). We assume an COVID-19 ICU capacity of 155 patients and that, starting September 1st, a 45% efficacious influenza vaccine is rolled out by age group proportional to historical trends (Figure S7), scaled to achieve 0%, 30%, or 60% coverage (Table S2).

Across all scenarios, COVID-19 is projected to dwarf seasonal influenza, in terms of incidence (Figure 1), ICU demand, and mortality (Figure S1). If non-pharmacological measures suppress COVID-19 and influenza transmission to low levels, then influenza vaccination rates are expected to only minimally ameliorate disease burden. Under the most pessimistic (high transmission) scenario, achieving high influenza vaccination coverage (60%) relative to low coverage (30%) is expected to reduce influenza-related illness, hospitalizations, ICU cases, and deaths by 17%, 18%, 18% and 22%, respectively (Figures 1, S1-S4). We project that ICU’s will be overwhelmed by COVID-19 patients in all but the lowest transmission scenarios, and that even high levels of influenza vaccination would not ensure sufficient capacity (Figure S1).

**Figure 1.**
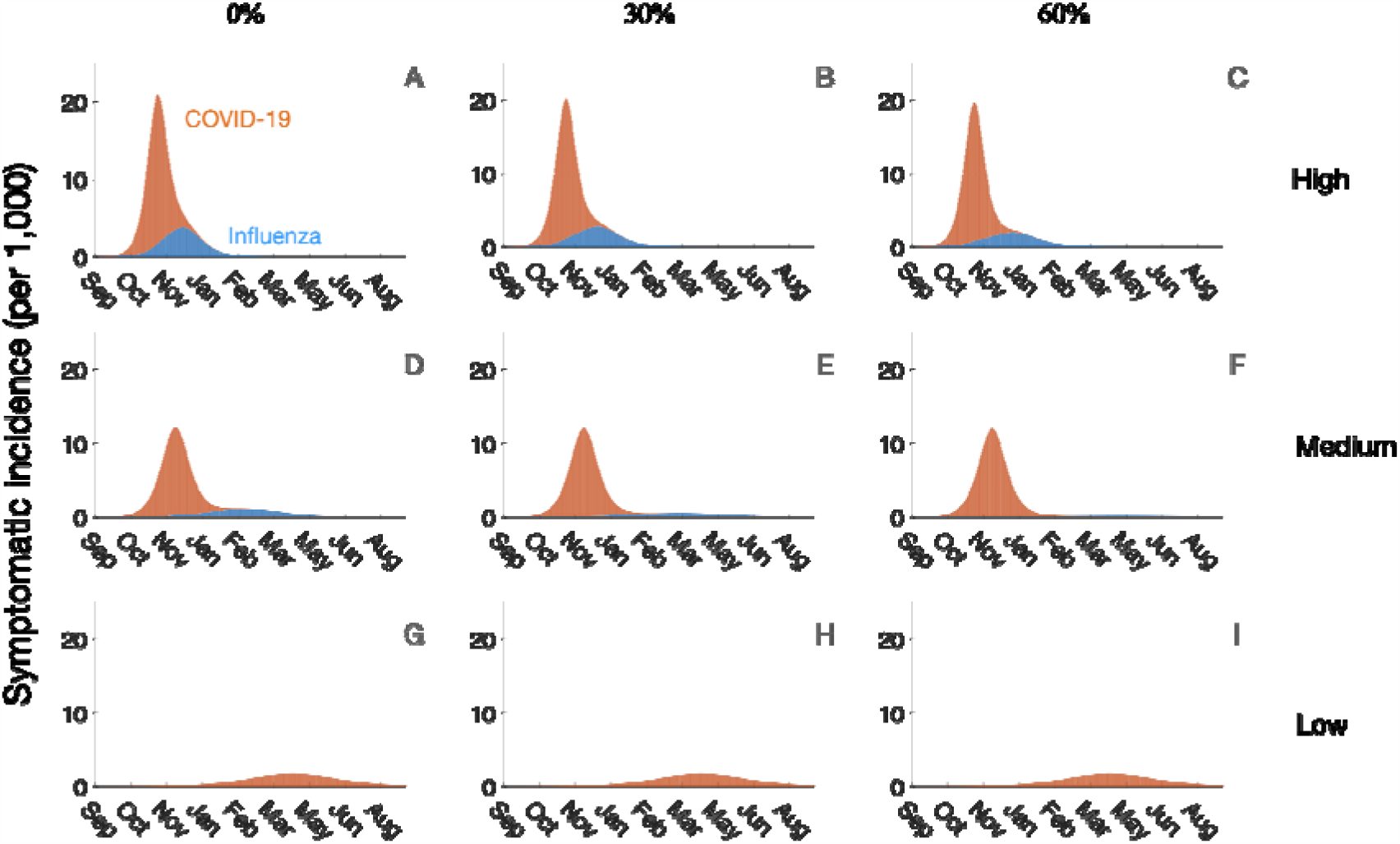
Projected incidence of symptomatic COVID-19 and influenza depending on the viral transmission rates (rows) resulting from COVID-19 community mitigation efforts and influenza vaccination coverage (columns), September 1, 2020 - Aug. 31, 2021. Using a deterministic model of the co-circulation of influenza and SARS-CoV-2 in Austin, Texas (Travis County) with the parameters given Tables S1 and S3, we consider three transmission scenarios: high (A-C), medium (E-F), and low (G-I), and three levels of influenza vaccination coverage: 0% (A, D, G), 30% (B, E, H) to 60% (D, F, I). The stacked curves indicate the combined daily incidence of symptomatic COVID-19 (orange) and influenza (blue) cases. The estimated COVID-19 and influenza related mortality and hospitalizations for each of the nine scenarios are given in Table S2.

COVID-19 will likely strain healthcare systems throughout the 2020-2021 influenza season. In communities that succeed in suppressing the spread of both viruses (1,2), influenza vaccination would be expected to only slightly reduce the burden of viral illness. If the viruses spread widely, influenza vaccination could prevent large numbers of cases, hospitalizations and deaths, as well as reduce laboratory burden and clinical uncertainty. Under all scenarios, COVID-19 hospitalizations would likely be orders of magnitude greater than influenza hospitalizations. Extrapolating from our projections, even universal influenza vaccine uptake would barely relieve ICU capacity.

As a caveats, we note that not all COVID-19 community mitigation measures will equally impact influenza. In particular, mass surveillance testing for COVID-19 to facilitate the safe opening of schools, workplaces and other public venues may prevent COVID-19 but not influenza spread. In addition, we assume that there is no cross-immunity between the viruses(4); competitive interactions might reduce the burden or slow the transmission of one or both viruses. Finally, the COVID-19 pandemic has reduced access to preventative care(5) and may thereby reduce influenza vaccination rates. Low levels of naturally- and vaccine-acquired immunity in 2021-2022 may increase the risk for more severe 2021-2022 seasonal influenza epidemics.

Mitigation of COVID-19 should take precedence over influenza vaccination, particularly in communities struggling to combat alarming healthcare surges. Nonetheless, aggressive influenza vaccine campaigns are critical for reducing morbidity and mortality in communities that are unable to sustain strict social distancing and face mask measures, for ensuring resilience towards future influenza seasons, and breaking down hesitancy towards future COVID-19 vaccination campaigns.

## Data Availability

Data available within the article or its supplementary materials.

## Acknowledgements

This work was supported by grant U01IP001136 from the Centers for Disease Control, Tito’s Handmade Vodka, and the Society for Medical Decision Making (SMDM) COVID-19 Decision Modeling Initiative (UTA20-000825). The authors acknowledge the Texas Advanced Computing Center (TACC) at The University of Texas at Austin for providing HPC, visualization, database, and grid resources that have contributed to the research results reported within this paper. URL: http://www.tacc.utexas.edu. The funders had no role in the design and conduct of the study; collection, management, analysis, and interpretation of the data; preparation, review, or approval of the manuscript; or decision to submit the manuscript for publication.

## References

1. Olsen SJ. Decreased Influenza Activity During the COVID-19 Pandemic — United States, Australia, Chile, and South Africa, 2020. MMWR Morb Mortal Wkly Rep [Internet]. 2020 [cited 2020 Nov 13];69. Available from: https://www.cdc.gov/mmwr/volumes/69/wr/mm6937a6.htm

2. CDC. COVIDView: A Weekly Surveillance Summary of U.S. COVID-19 Activity [Internet]. 2020 [cited 2020 Nov 15]. Available from: https://www.cdc.gov/coronavirus/2019-ncov/covid-data/covidview/index.html

3. Wang X, Pasco RF, D. Z, Petty M, Fox SJ, Galvani AP, et al. Impact of Social Distancing Measures on Coronavirus Disease Healthcare Demand, Central Texas, USA. Emerg Infect Dis. 2020 Oct;26(10):2361–9.

4. Bolourian A, Mojtahedi Z. COVID-19 and Flu Pandemics Follow a Pattern: A Possible Cross-immunity in the Pandemic Origin and Graver Disease in Farther Regions. Arch Med Res [Internet]. 2020 Oct 17; Available from: http://www.sciencedirect.com/science/article/pii/S018844092031609X

5. NFID Call to Action: Dangers of Influenza and COVID-19 in Adults with Chronic Health Conditions.pdf. Available from: https://www.nfid.org/wp-content/uploads/2020/10/NFID-Call-to-Action-Dangers-of-Influenza-and-COVID-19-in-Adults-with-Chronic-Health-Conditions.pdf

