## Appendix for "Projecting the combined healthcare burden of seasonal influenza and COVID-19"

**Supplement**

##### **Table S1. Key parameters for the SEIR compartmental model of COVID-19 and influenza co-circulation and nine transmission-vaccination scenarios.**

|  | **Transmission scenario** | | | | | |
| --- | --- | --- | --- | --- | --- | --- |
|  | **COVID-19** | | | **Influenza** | | |
| ***R*_0_** | 2.20(1) | | | 2.05 (2) | | |
| **Prior Immunity** | 6% (3) | | | 35% (2) | | |
|  | **High** | | **Medium** | | **Low** | |
|  | **COVID-19** | **Influenza** | **COVID-19** | **Influenza** | **COVID-19** | **Influenza** |
| **Mitigation (**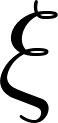<https://www.codecogs.com/eqnedit.php?latex=%5Cxi#0>**)** | 0% | | 13% | | 58% | |
| ***R_e_*^⇞^** | 2.07**^+^** | 1.33**^+^** | 1.80(3) | 1.16**^+^** | 1.2(3) | 0.77**^+^** |
| 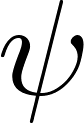<https://www.codecogs.com/eqnedit.php?latex=%5Cpsi#0>**: influenza vaccine coverage** | 0%, 30%, and 60% | | | | | |
| 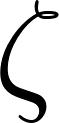<https://www.codecogs.com/eqnedit.php?latex=%5Czeta#0>**: influenza efficacy** | 45% (4) | | | | | |
| **ICU capacity**  **(Travis county)** | 155 (5) | | | | | |

**^⇞^** The effective reproduction numbers (*R_e_*) for each scenario is derived by reducing the basic reproduction numbers (*R_0_*) to account for both prior immunity and the impacts of non-pharmacological interventions, as given by *R_e_* = *R_0_* (1-
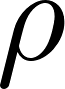
<https://www.codecogs.com/eqnedit.php?latex=%5Crho#0>)(1-
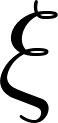
<https://www.codecogs.com/eqnedit.php?latex=%5Cxi#0>) where
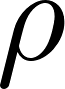
<https://www.codecogs.com/eqnedit.php?latex=%5Crho#0> is the prior immunity in the population and
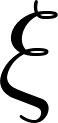
<https://www.codecogs.com/eqnedit.php?latex=%5Cxi#0> is the impact of mitigation. The *high transmission* scenario assumes no mitigation; the *low* and *medium* scenarios are based on the estimated COVID-19 transmission reductions in Austin, Texas in mid-May and mid-June of 2020, respectively (3).

##### **Table S2. Projected cumulative mortality and hospitalizations under the nine transmission-vaccination scenarios. Death and daily admissions, from September 1, 2020 to August 31, 2021.**

| **Scenarios** | | | **Outcomes** | |
| --- | --- | --- | --- | --- |
| ***R_e_* of COVID-19** | ***R_e_* of influenza** | **Influenza** **vaccination coverage** | **Total deaths per 100,000** | **Total hospitalization per 100,000** |
| 2.2 | 1.44 | 0% | 105 | 4710 |
| 2.2 | 1.44 | 30% | 104 | 4640 |
| 2.2 | 1.44 | 60% | 103 | 4586 |
| 1.8 | 1.18 | 0% | 84 | 3891 |
| 1.8 | 1.18 | 30% | 84 | 3841 |
| 1.8 | 1.18 | 60% | 83 | 3797 |
| 1.2 | 0.79 | 0% | 32 | 1607 |
| 1.2 | 0.79 | 30% | 32 | 1607 |
| 1.2 | 0.79 | 60% | 32 | 1607 |

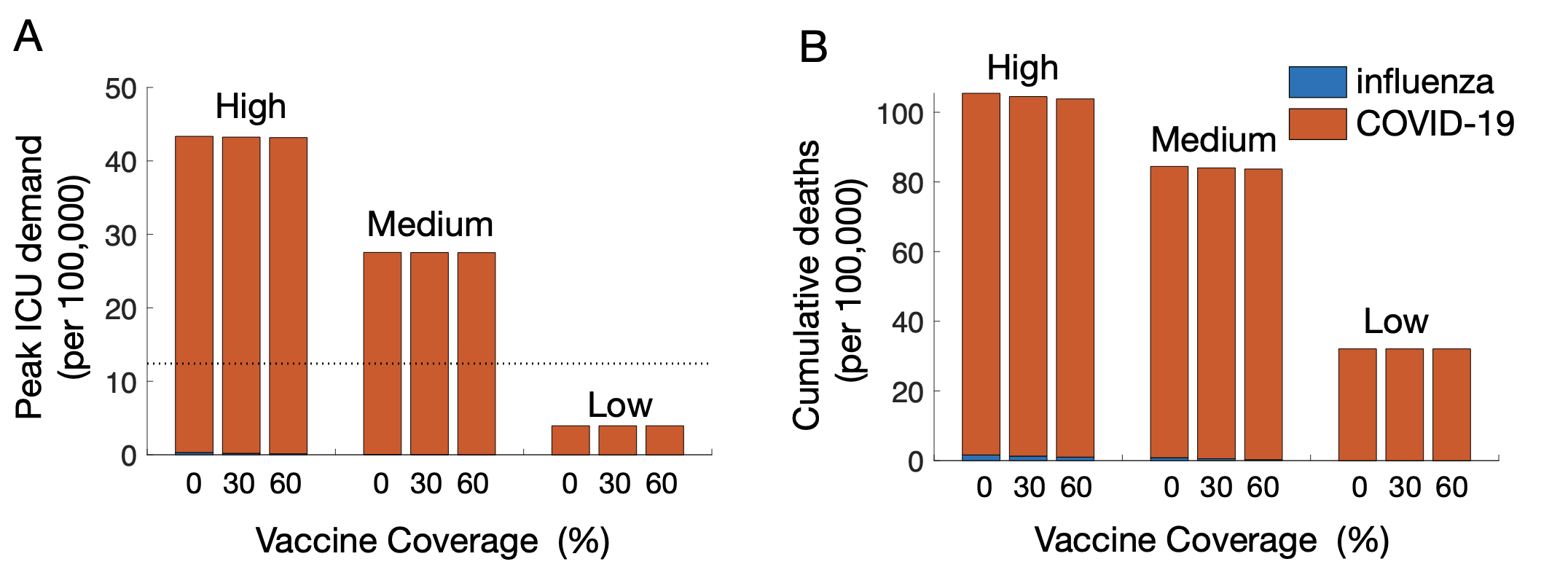

**Figure S1. Projected COVID-19 and influenza burden depending on non-pharmacological interventions and influenza vaccination coverage in Austin, Texas (Travis County) from September 1, 2020 through Aug. 31, 2021.** Estimates are derived using a deterministic model with parameters in Table S3 for all combinations of three levels of transmission rates (high, medium and low) and three influenza vaccination rates (0%, 30% and 60%). (A) Peak number of ICU patients. The dotted line indicates the COVID-19 ICU capacity estimated Travis County of 155 patients (5). (B) Cumulative deaths during the projected period.

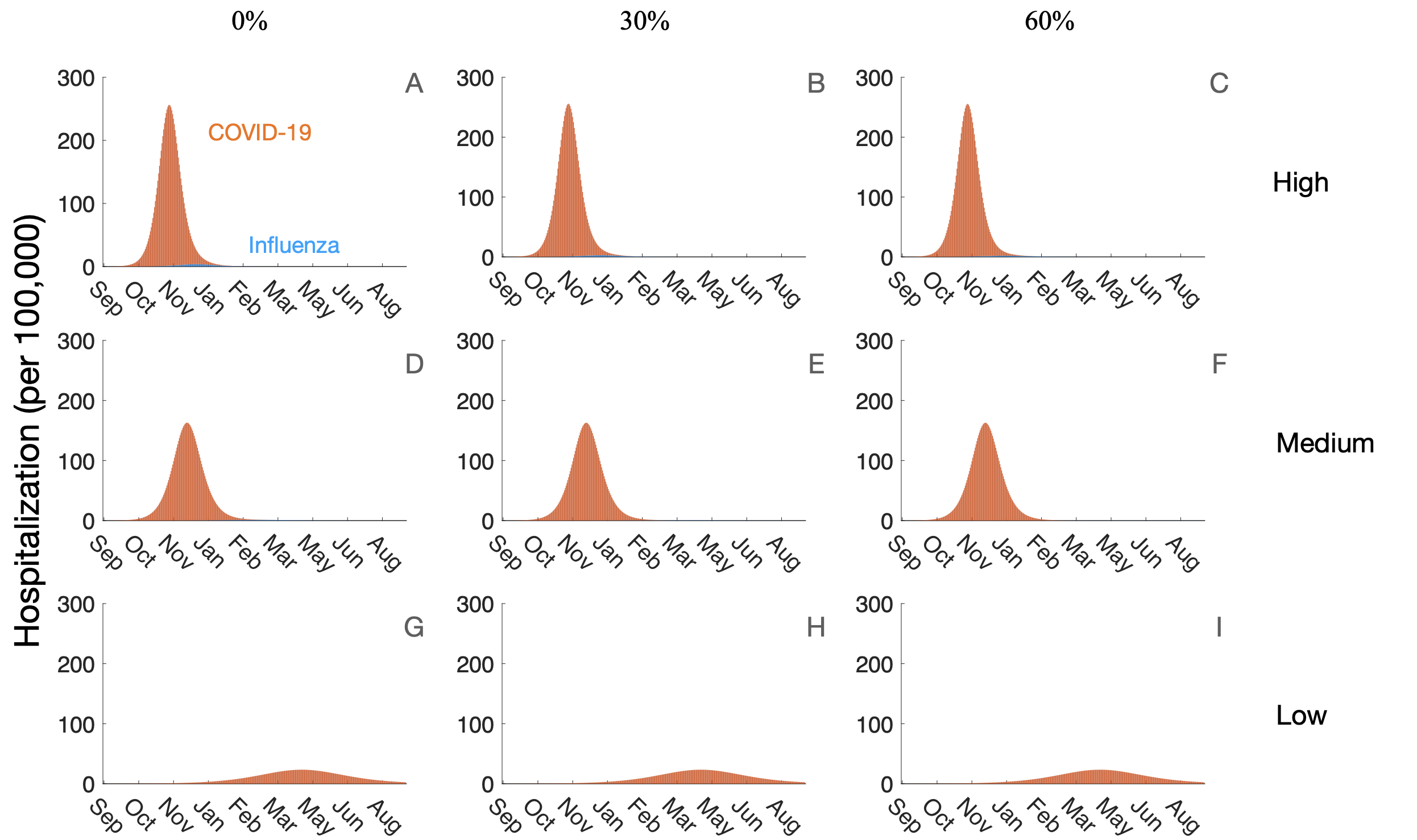

**Figure S****2. Projected COVID-19 and influenza hospitalizations depending on the viral transmission rates (rows) resulting from COVID-19 community mitigation efforts and influenza vaccination coverage (columns), September 1, 2020 - Aug. 31, 2021.** Using a deterministic model of the co-circulation of influenza and SARS-CoV-2 in Austin, Texas (Travis County) with the parameters given Tables S1 and S3, we consider three transmission scenarios: high (A-C), medium (E-F), and low (G-I), and three levels of influenza vaccination coverage: 0% (A, D, G), 30% (B, E, H) to 60% (D, F, I). The stacked curves indicate the combined daily hospitalizations due to COVID-19 (orange) and influenza (blue).

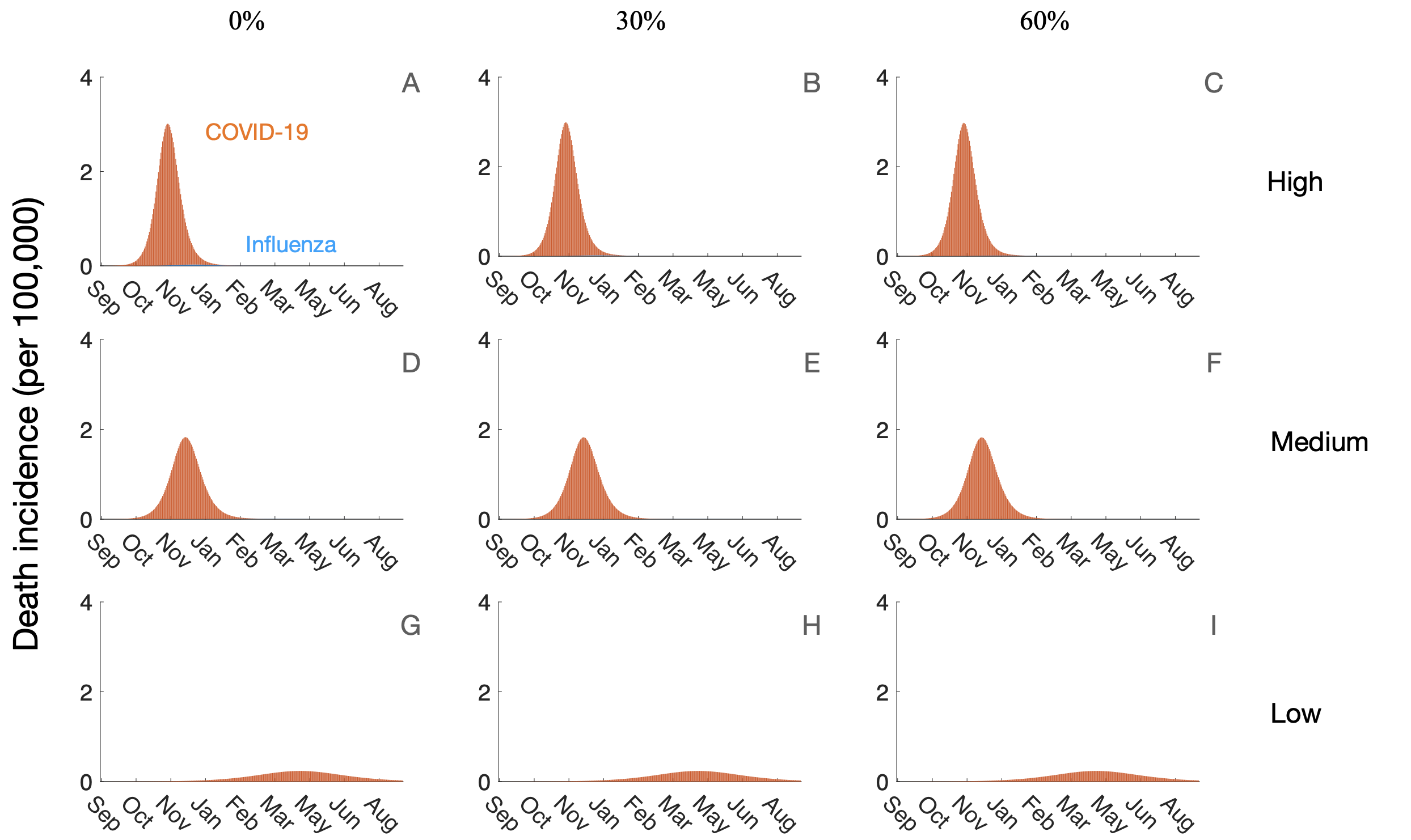

**Figure S****3. Projected COVID-19 and influenza mortality depending on the viral transmission rates (rows) resulting from COVID-19 community mitigation efforts and influenza vaccination coverage (columns), September 1, 2020 - Aug. 31, 2021.** Using a deterministic model of the co-circulation of influenza and SARS-CoV-2 in Austin, Texas (Travis County) with the parameters given Tables S1 and S3, we consider three transmission scenarios: high (A-C), medium (E-F), and low (G-I), and three levels of influenza vaccination coverage: 0% (A, D, G), 30% (B, E, H) to 60% (D, F, I). The stacked curves indicate the combined daily death incidence due to COVID-19 (orange) and influenza (blue).

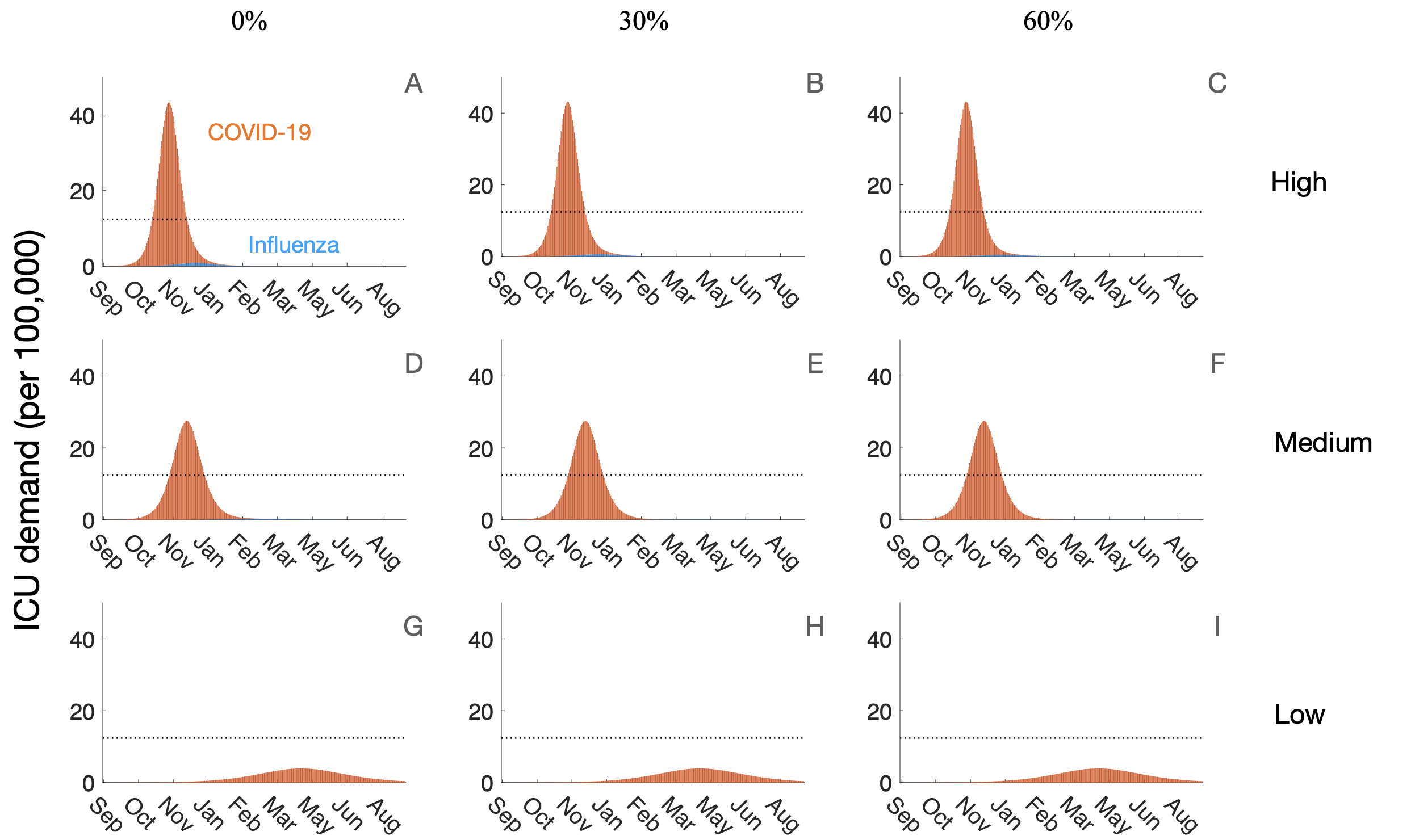

**Figure S****4. Projected COVID-19 and influenza ICU demand depending on the viral transmission rates (rows) resulting from COVID-19 community mitigation efforts and influenza vaccination coverage (columns), September 1, 2020 - Aug. 31, 2021.** Using a deterministic model of the co-circulation of influenza and SARS-CoV-2 in Austin, Texas (Travis County) with the parameters given Tables S1 and S3, we consider three transmission scenarios: high (A-C), medium (E-F), and low (G-I), and three levels of influenza vaccination coverage: 0% (A, D, G), 30% (B, E, H) to 60% (D, F, I). The stacked curves indicate the combined daily hospitalizations due to COVID-19 (orange) and influenza (blue). The dotted line indicates the COVID-19 ICU capacity estimated Travis County of 155 patients (5).

##### **Table S3. General epidemiological parameters for the COVID-19 and influenza co-spreading infection dynamic model.** Values given as five-element vectors are age-stratified with values corresponding to 0-4, 5-17, 18-49, 50-64, 65+ year age groups, respectively.

| **Parameters** | **Values** | |
| --- | --- | --- |
|  | **Influenza** | **COVID-19** |
| Initial day of simulation | Sept. 1, 2020 | |
| Population in Travis county in 2018, age specific | [78395, 192140, 651139, 203635, 123434] (6) | |
| Initial infection number | 100 cases | 100 cases |
| 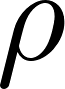<https://www.codecogs.com/eqnedit.php?latex=%5Crho#0>: prior immunity (proportion of the population already immune, as of September 1, 2020) | 35% (2) | 6% (7) |
| 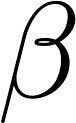<https://www.codecogs.com/eqnedit.php?latex=%5Cbeta#0>: transmission rate | Calibrated to specified reproduction number via next generation matrix method (8) (See Appendix 1) | |
| 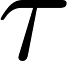<https://www.codecogs.com/eqnedit.php?latex=%5Ctau#0>: symptomatic proportion | 0.55 (9) | 0.75 (10) |
| 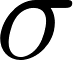<https://www.codecogs.com/eqnedit.php?latex=%5Csigma#0>: exposed rate at which individuals progress from E to P | 1/0.98 (11) | 1/3 (12) |
| 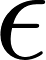<https://www.codecogs.com/eqnedit.php?latex=%5Cepsilon#0>: rate at which individuals progress from P to Y | 2 (13) | 1/2 (12) |
| 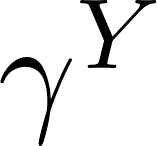<https://www.codecogs.com/eqnedit.php?latex=%5Cgamma%5EY#0>: recovery rate of symptomatic cases from Y to R | 1/5 (11) | 1/4 (10,14) |
| 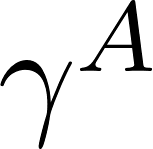<https://www.codecogs.com/eqnedit.php?latex=%5Cgamma%5EA#0>: recovery rate of symptomatic cases from A to R | 1/3 (13) | 1/9 (15) |
| 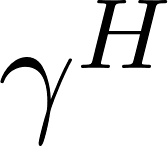<https://www.codecogs.com/eqnedit.php?latex=%5Cgamma%5EH#0>: recovery rate in hospitalized compartment | 1 (11,16) | 1/2 (10) |
| 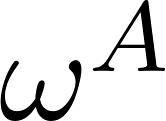<https://www.codecogs.com/eqnedit.php?latex=%5Comega%5EA#0>: relative infectiousness of infectious individuals in compartment A | 0.36 (13) | 0.5 (12) |
| 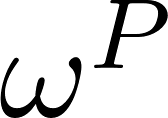<https://www.codecogs.com/eqnedit.php?latex=%5Comega%5EP#0>: relative infectiousness of infectious individuals in compartment P | 0.68 (13) | 1.57 (10,14) |
| 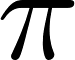<https://www.codecogs.com/eqnedit.php?latex=%5Cpi#0>: rate at which individuals progress from Y to H | 1/4.6 (16) | 1/2 (10) |
| *h*: hospitalization ratio, age specific (%) | [0.7, 0.27, 0.56, 1.1, 9.09] (17) | [0, 0.025, 2.672, 9.334, 15.465] (18) |
| 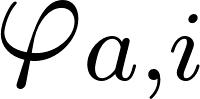<https://www.codecogs.com/eqnedit.php?latex=%5Cvarphi_%7Ba%2Ci%7D#0>: contact rate between individuals of age *a* and *i* | Mean contact rates estimated within and between age groups (19) | |
| *e*: death rate of hospitalization, age specific (%) | [0.0050, 0.0072, 0.040, 0.079, 1.57] (17) | [0.0016, 0.0049, 0.084, 1.000, 3.371] (20) |
| proportion hospitalized people in ICU, age specific | [0.14, 0.20, 0.23, 0.24, 0.24] (21) | [0.15, 0.20, 0.15, 0.20, 0.15] (21) |

##### **Table S4. Parameters for age-structured influenza vaccine roll-out model.** Values given as five-element vectors corresponding to *b*, *c*, *d*, *e*, and *f* in the log-logistic function of the form <https://www.codecogs.com/eqnedit.php?latex=%5Cdelta_%7Ba%2Ct%7D%20%3Dc%2B%5Cfrac%7Bd-c%7D%7B((1%20%2B%20exp(b(log(t)%20-%20log(e))))%5Ef%7D#0>where <https://www.codecogs.com/eqnedit.php?latex=%5Cdelta_%7Ba%2Ct%7D%20#0> is the cumulative influenza vaccination coverage of age group *a* at time *t* and *b*, *c*, *d*, *e*, and *f* are parameter values unique to each age group (Figure S6).

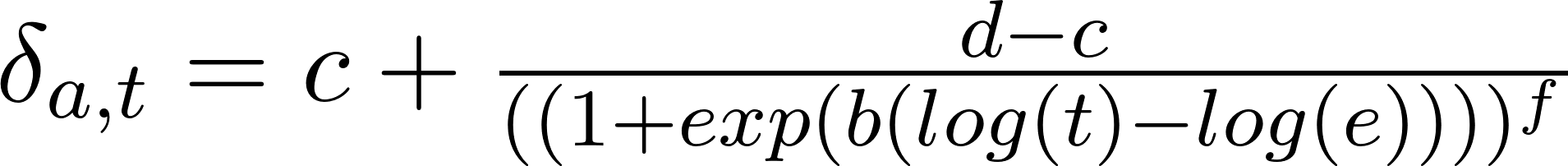

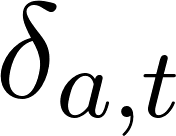

| **Age Cohort** | **Parameters for Vaccine Roll-Out Function** |
| --- | --- |
| 6 months - 4 years | [-0.02676, -8.576, 70.45, 18520, 1.037] |
| 5 years - 17 years | [-0.03007, 0.700, 56.60, 18490, 2.402] |
| 18 years - 49 years | [-0.02463, -5.686, 30.51, 18510, 1.327] |
| 50 years - 64 years | [-0.03218, -1.793, 42.42, 18480, 4.002] |
| 65+ years | [-0.03459, -14.56, 62.10, 18510, 1.054] |

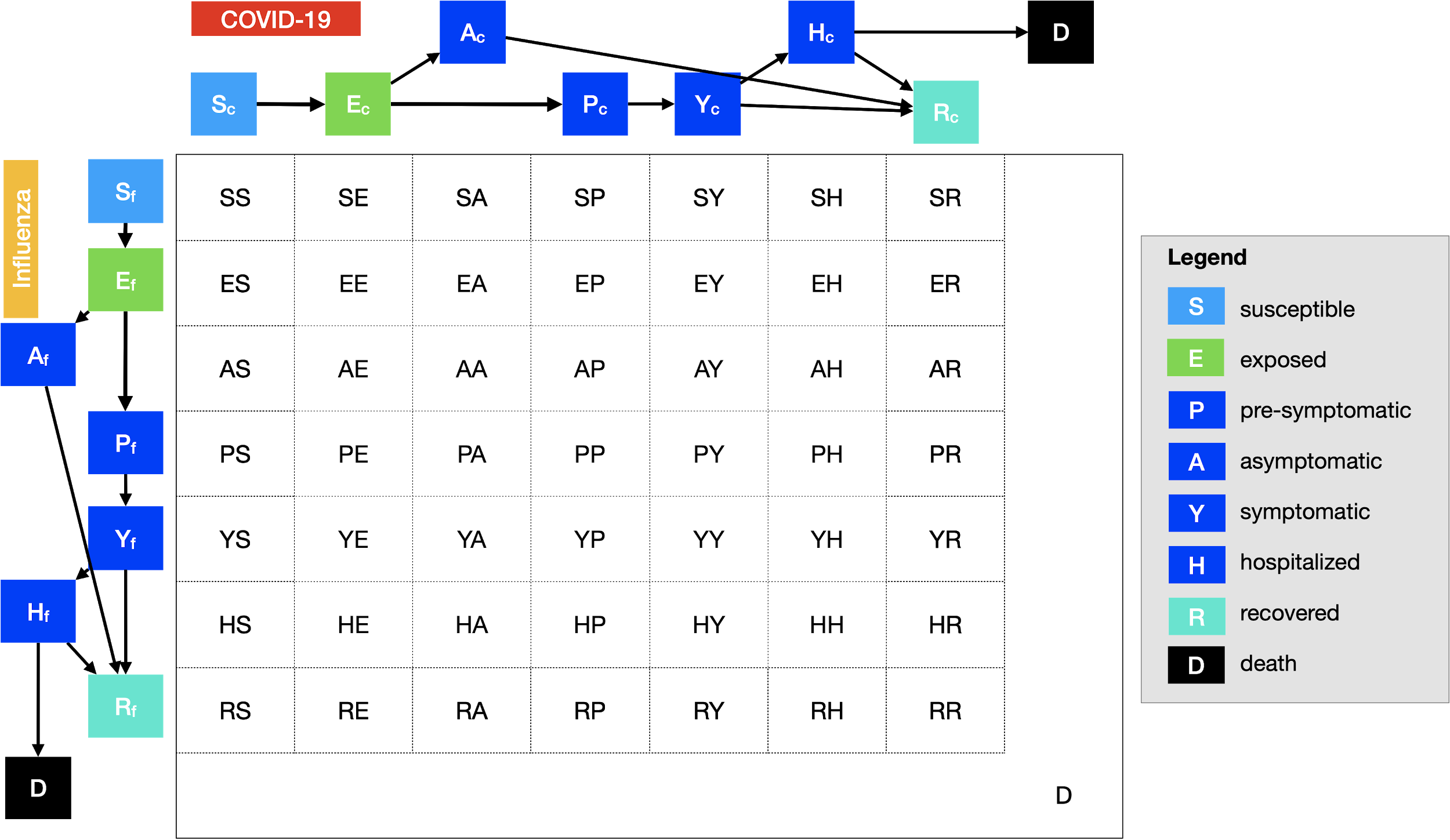

**Figure S****5. Compartmental model of COVID-19 and influenza transmission.** Each of five age groups is modeled with a separate set of 50 compartments. Upon infection by either virus, susceptible individuals (*S*) progress to exposed (*E*). Exposed individuals progress to either pre-symptomatic infectious (*P*) or asymptomatic infectious (*A*). Pre-symptomatic individuals will progress to symptomatic (*Y*) and then to either hospitalized (*H*) or recover (*R*). All asymptomatic cases progress to recover (*R*). Hospitalized cases either recover (*R*) or die (*D*). To model vaccination, we move individuals from *S* to *R* based on age-specific daily vaccination rates and vaccine efficacy.

**Appendix 1. Epidemic Model**

The structure of the deterministic susceptible-exposed-infected-recovered compartmental model of SARS-CoV-2 and influenza co-circulation is diagrammed in Figure S5 and described in the equations below.

For each of the five age groups, we build a separate set of compartments to model the transitions between the states for each disease: susceptible (*S*), exposed (*E*), asymptomatic (A), pre-symptomatic (*P*), symptomatic (*Y*), symptomatic infectious that are hospitalized (*H*), recovered (*R*), and deceased (*D*). The symbols *S, E, A, P, Y, H, R*, and *D* denote the number of people in that state in the given age group. We deonte COVID-19 and influenza as *c* and *f* in subscripts, respectively. The model for individuals in age group *a* and virus *v* (either influenza or COVID-19) is given by:

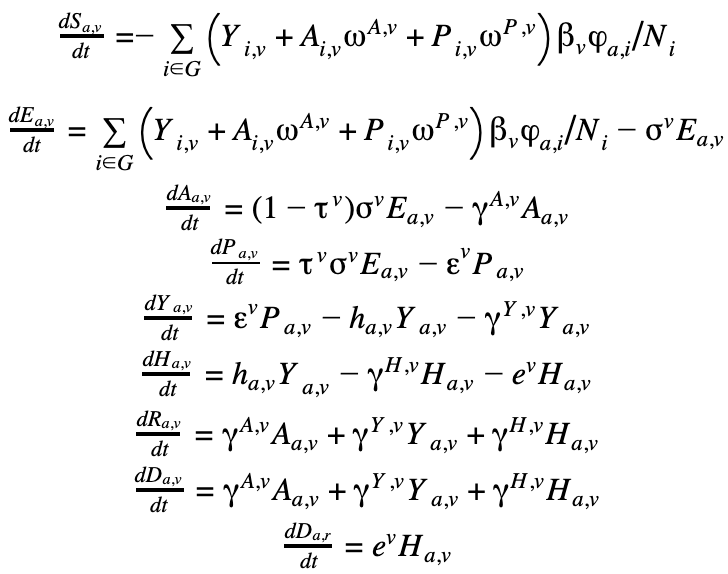

where *G* are the five age groups,
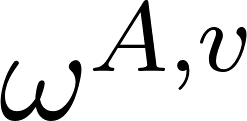
<https://www.codecogs.com/eqnedit.php?latex=%5Comega%5E%7BA%2Cv%7D#0>,
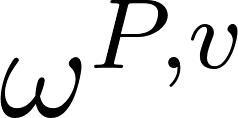
<https://www.codecogs.com/eqnedit.php?latex=%5Comega%5E%7BP%2Cv%7D#0> are relative infectiousness of the *A* and *P* compartments of virus *v*, respectively, and
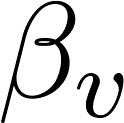
<https://www.codecogs.com/eqnedit.php?latex=%5Cbeta_v#0> is the transmission rate of the virus.

Parameter details and values are provided in Tables S1 and S3. For each transmission scenario (low, medium and high), we reduced published estimates for the basic reproduction number (*R*_0_) to account for mitigation. Then, we use the next generation matrix approach to solve for the corresponding transmission rate (8). Simulations are initiated assuming 94% and 65% of all age groups are susceptible to COVID-19 and influenza, respectively, based on the estimated cumulative infections in Austin-Round Rock MSA through Sept. 1, 2020 (3) and the median susceptibility rate of influenza seasons from 2004 to 2013 (2).

Let
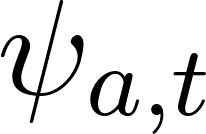
<https://www.codecogs.com/eqnedit.php?latex=%5Cpsi_%7Ba%2Ct%7D#0> denote the daily influenza vaccination coverage of age group *a* at time *t* and
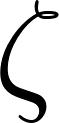
<https://www.codecogs.com/eqnedit.php?latex=%5Czeta#0> denote the influenza vaccination efficacy. For each age group *a* at time *t*, we move a fraction
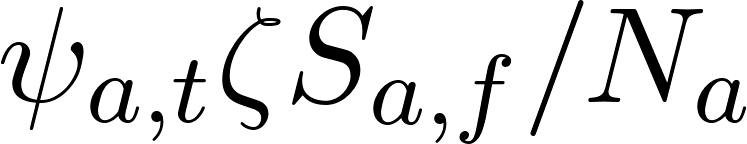
<https://www.codecogs.com/eqnedit.php?latex=%5Cpsi_%7Ba%2Ct%7D%5Czeta%20S_%7Ba%2Cf%7D%2FN_a#0> of each influenza-susceptible class (*S_a,f_*) to a influenza-recovered class (*R_a,f_*), while preserving COVID-19 state. Here *N_a_* is the number of individuals in age group *a*.

Although we focus our estimates on Austin, Texas (Travis County), we expect our projections to apply broadly to US cities, with some caveats. COVID-19 transmission will depend on population-wide immunity accumulated through the initial months of the pandemic. Regions like New York City, where a quarter of the population may have been infected by September 2020 (22), may experience smaller or slower winter COVID-19 epidemics than regions with lower levels of population immunity. The spread of both viruses will depend on community compliance with mitigation orders, which vary both geographically and temporally as policies and opinions change. The healthcare impacts will also depend on local hospital capacity and the socioeconomic and demographic composition of the community.

**Appendix 2. Vaccination Coverage Estimation**

The daily age-specific vaccination rates are based on monthly flu vaccine coverage data curated by the CDC for the state of Texas over recent flu seasons from 2014 to 2018 (23). To determine daily vaccine coverage, we fit a log-logistic curve for each of the following age groups (6 months - 4 years, 5-17 years, 18-49 years, 50-64 years, and 65+ years)(24). The log-logistic curves are of the form
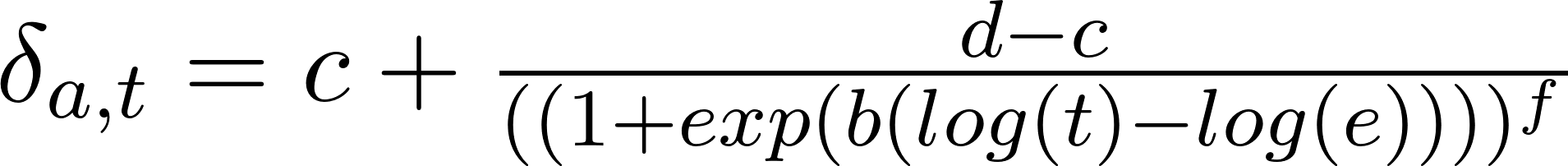
<https://www.codecogs.com/eqnedit.php?latex=%5Cdelta_%7Ba%2Ct%7D%20%3Dc%2B%5Cfrac%7Bd-c%7D%7B((1%20%2B%20exp(b(log(t)%20-%20log(e))))%5Ef%7D#0>where
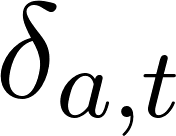
<https://www.codecogs.com/eqnedit.php?latex=%5Cdelta_%7Ba%2Ct%7D%20#0> is the cumulative influenza vaccination coverage of age group *a* at time *t* , and *b*, *c*, *d*, *e*, and *f* are parameters unique to each age group, as described in **Table S4** and depicted in Figure S6. We derive the daily influenza vaccination roll-out to age group *a* at time *t* (
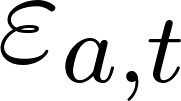
<https://www.codecogs.com/eqnedit.php?latex=%5Cvarepsilon_%7Ba%2Ct%7D%20#0>), from
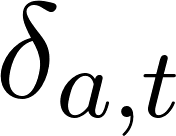
<https://www.codecogs.com/eqnedit.php?latex=%5Cdelta_%7Ba%2Ct%7D%20#0>. For each vaccination coverage scenario (
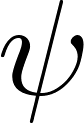
<https://www.codecogs.com/eqnedit.php?latex=%5Cpsi#0>) in **Table S1**, we assume that the number of individuals in age group *a* vaccinated at time *t* (
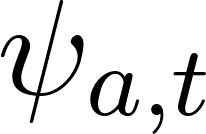
<https://www.codecogs.com/eqnedit.php?latex=%5Cpsi_%7Ba%2Ct%7D#0>) is
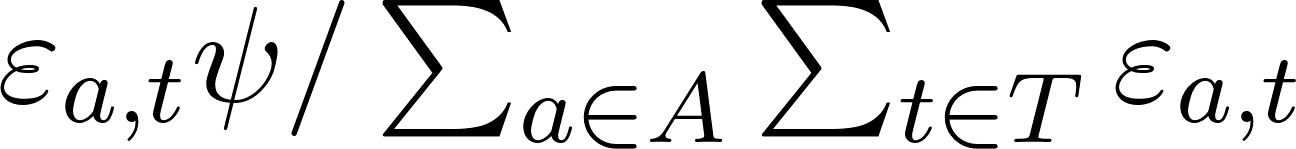
<https://www.codecogs.com/eqnedit.php?latex=%5Cvarepsilon_%7Ba%2Ct%7D%5Cpsi%20%2F%20%5Csum_%7Ba%20%5Cin%20A%7D%20%5Csum_%7Bt%20%5Cin%20T%7D%20%5Cvarepsilon_%7Ba%2Ct%7D%20%20#0> (**Figure S7**).

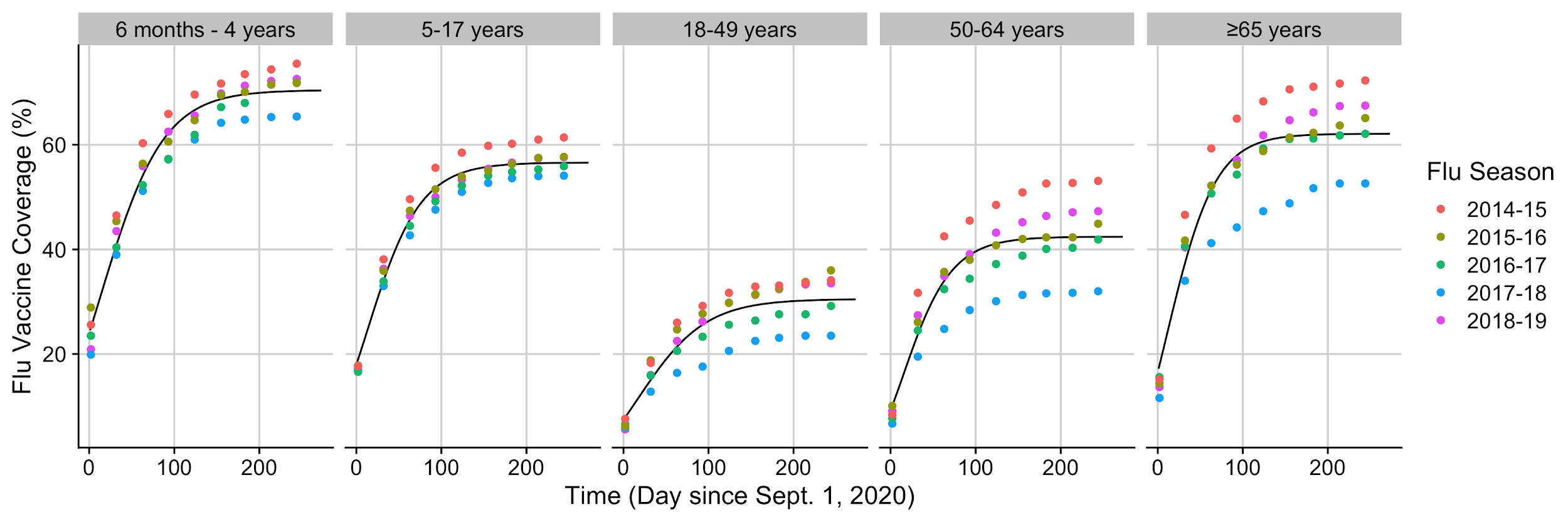

**Figure S****6.** **Age-stratified roll-out of seasonal influenza vaccines in the US.** We fit log-logistic functions (black curves) to the reported vaccine coverage data of Texas over recent flu seasons, from 2014-2015 to 2018-2019(23). The fitted model predicts an average population coverage of 42%.

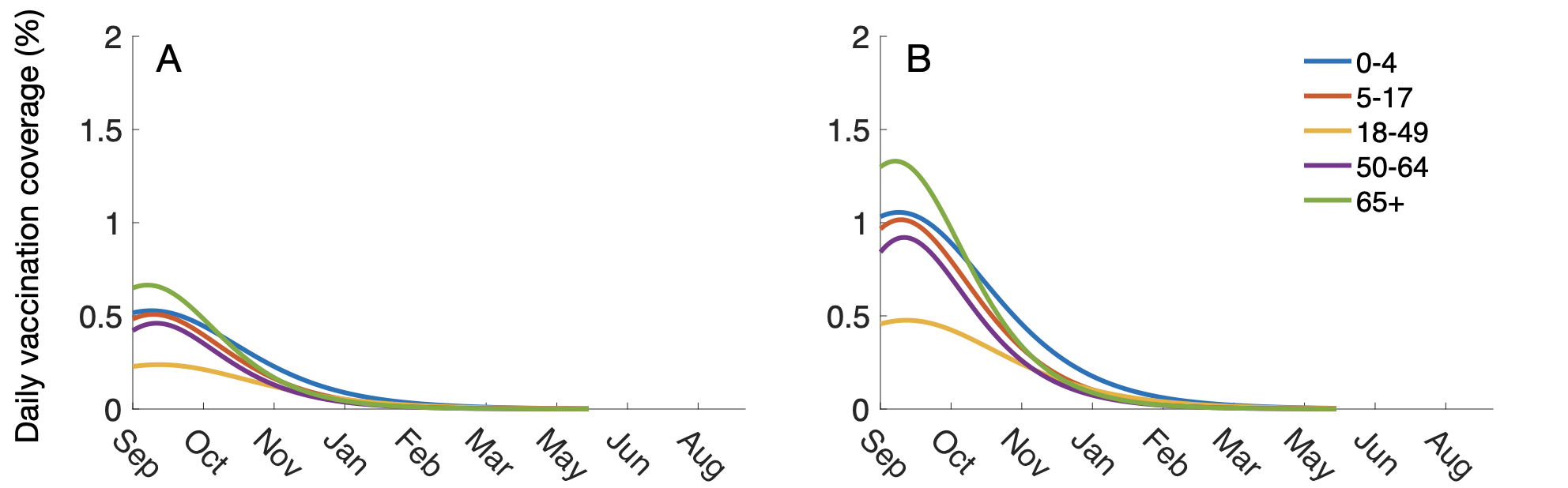

**Figure S****7. Age-stratified seasonal influenza vaccination timelines for the (A) 30% and (B) 60% coverage scenarios, daily from September 1, 2020 through August 31, 2021.**

**References**

1.	Li Q, Guan X, Wu P, Wang X, Zhou L, Tong Y, et al. Early Transmission Dynamics in Wuhan, China, of Novel Coronavirus-Infected Pneumonia. N Engl J Med. 2020 Mar 26;382(13):1199–207.

2.	Yang W, Lipsitch M, Shaman J. Inference of seasonal and pandemic influenza transmission dynamics. Proc Natl Acad Sci U S A. 2015 Mar 3;112(9):2723–8.

3.	Lachmann M, Fox SJ, Tec M, Pasco R, Du Z, Woody S, et al. Texas Trauma Service Area (TSA) COVID-19 transmission estimates and healthcare projections: Oct. 20 Update. 2020;

4.	[Dawood FS, Chung JR, Kim SS, Zimmerman RK, Nowalk MP, Jackson ML, et al. Interim Estimates of 2019–20 Seasonal Influenza Vaccine Effectiveness — United States, February 2020 [Internet]. Vol. 69, MMWR. Morbidity and Mortality Weekly Report. 2020. p. 177–82. Available from:](http://paperpile.com/b/q7QcPE/8FPxL) <http://dx.doi.org/10.15585/mmwr.mm6907a1>

5.	[America’s COVID warning system [Internet]. [cited 2020 Oct 31]. Available from:](http://paperpile.com/b/q7QcPE/D6nfV) <https://covidactnow.org/us/texas-tx/county/travis_county?s=1226301>

6.	[US Census Bureau. 2018 American Community Survey Single-Year Estimates [Internet]. [cited 2020 Oct 24]. Available from:](http://paperpile.com/b/q7QcPE/j6g7M) <https://www.census.gov/newsroom/press-kits/2019/acs-1year.html>

7.	Tec M, Lachmann M, Fox SJ, Pasco R, Woody S, Starling J, et al. Austin COVID-19 transmission estimates and healthcare projections. Available from: <https://sites.cns.utexas.edu/sites/default/files/cid/files/austin_dashboard_report_071520.pdf>

8.	Diekmann O, Heesterbeek JAP, Roberts MG. The construction of next-generation matrices for compartmental epidemic models. J R Soc Interface. 2010 Jun 6;7(47):873–85.

9.	Leung NHL, Xu C, Ip DKM, Cowling BJ. Review Article: The Fraction of Influenza Virus Infections That Are Asymptomatic: A Systematic Review and Meta-analysis. Epidemiology. 2015 Nov;26(6):862–72.

10.	[Aleta A, Martín-Corral D, Pastore Y Piontti A, Ajelli M, Litvinova M, Chinazzi M, et al. Modelling the impact of testing, contact tracing and household quarantine on second waves of COVID-19. Nat Hum Behav [Internet]. 2020 Aug 5; Available from:](http://paperpile.com/b/q7QcPE/6BD95) <http://dx.doi.org/10.1038/s41562-020-0931-9>

11.	Yang Y, Sugimoto JD, Halloran ME, Basta NE, Chao DL, Matrajt L, et al. The transmissibility and control of pandemic influenza A (H1N1) virus. Science. 2009 Oct 30;326(5953):729–33.

12.	Backer JA, Klinkenberg D, Wallinga J. Incubation period of 2019 novel coronavirus (2019-nCoV) infections among travellers from Wuhan, China, 20-28 January 2020. Euro Surveill. 2020 Feb;25(5):2000062.

13.	Wu JT, Leung GM, Lipsitch M, Cooper BS, Riley S. Hedging against antiviral resistance during the next influenza pandemic using small stockpiles of an alternative chemotherapy. PLoS Med. 2009 May 19;6(5):e1000085.

14.	He X, Lau EHY, Wu P, Deng X, Wang J, Hao X, et al. Temporal dynamics in viral shedding and transmissibility of COVID-19. Nat Med. 2020 May;26(5):672–5.

15.	[Du Z, Pandey A, Bai Y, C. Fitzpatrick M, Chinazzi M, Pastore y Piontti A, et al. Comparative Cost-Effectiveness of SARS-CoV-2 Testing Strategies [Internet]. papers.ssrn.com. 2020 [cited 2020 Nov 14]. Available from:](http://paperpile.com/b/q7QcPE/CHAA2) <https://papers.ssrn.com/abstract=3714642>

16.	Biggerstaff M, Jhung MA, Reed C, Fry AM, Balluz L, Finelli L. Influenza-like illness, the time to seek healthcare, and influenza antiviral receipt during the 2010-2011 influenza season-United States. J Infect Dis. 2014 Aug 15;210(4):535–44.

17.	[CDC. Disease Burden of Influenza [Internet]. 2020 [cited 2020 Nov 14]. Available from:](http://paperpile.com/b/q7QcPE/DBwy5) <https://www.cdc.gov/flu/about/burden/index.html?CDC_AA_refVal=https%3A%2F%2Fwww.cdc.gov%2Fflu%2Fabout%2Fdisease%2Fburden.htm>

18.	[Aleta A, Martín-Corral D, Piontti APY, Ajelli M, Litvinova M, Chinazzi M, et al. Modeling the impact of social distancing, testing, contact tracing and household quarantine on second-wave scenarios of the COVID-19 epidemic. medRxiv [Internet]. 2020 May 18; Available from:](http://paperpile.com/b/q7QcPE/scuVF) <http://dx.doi.org/10.1101/2020.05.06.20092841>

19.	Prem K, Cook AR, Jit M. Projecting social contact matrices in 152 countries using contact surveys and demographic data. PLoS Comput Biol. 2017 Sep;13(9):e1005697.

20.	Verity R, Okell LC, Dorigatti I, Winskill P, Whittaker C, Imai N, et al. Estimates of the severity of coronavirus disease 2019: a model-based analysis. Lancet Infect Dis. 2020 Jun;20(6):669–77.

21.	Wang X, Pasco RF, Du Z, Petty M, Fox SJ, Galvani AP, et al. Impact of Social Distancing Measures on Coronavirus Disease Healthcare Demand, Central Texas, USA. Emerg Infect Dis. 2020 Oct;26(10):2361–9.

22.	[Goldstein J. 1.5 Million Antibody Tests Show What Parts of N.Y.C. Were Hit Hardest. The New York Times [Internet]. 2020 Aug 19 [cited 2020 Nov 15]; Available from:](http://paperpile.com/b/q7QcPE/qPF20) <https://www.nytimes.com/2020/08/19/nyregion/new-york-city-antibody-test.html>

23.	[Results for General Population Influenza Vaccination Coverage [Internet]. 2020 [cited 2020 Oct 26]. Available from:](http://paperpile.com/b/q7QcPE/KQOAp) <https://www.cdc.gov/flu/fluvaxview/interactive-general-population.htm>

24.	[Onofri A. The broken bridge between biologists and statisticians: a blog and R package [Internet]. Statforbiology; 2019. Available from:](http://paperpile.com/b/q7QcPE/buddP) <https://www.statforbiology.com/>
